# Environmental risks and occupational health hazards of bidi workers and their communities in India: protocol for systematic scoping review

**DOI:** 10.1101/2022.03.24.22272764

**Authors:** Jyoti Tyagi, Deepti Beri, Soumyadeep Bhaumik

**Author notes:** Correspondence to: Dr Soumyadeep Bhaumik.

## Abstract

**Background:** Tobacco consumption is a leading preventable cause of disease and premature deaths. In India bidis are the most common form of smoking tobacco product. Bidi manufacturing is an agro-forest based cottage industry and is generally rolled at home. These workers are exposed to nicotine, tar and other particles through skin leading to occupational health risks in not only themselves but also in their families and communities. There are concerns on environmental risks of bidi manufacturing in home too.

**Aim:** To assess the environmental risks and occupational health hazards of bidi workers and their communities in India.

**Method:** We will conduct a scoping review to identify, map and analyse evidence around environmental risks and occupational health hazards in bidi workers and their families and communities. We will search in seven electronic databases (PubMed, EMBASE+EMBASE Classic, CINAHL, Environment complete-EBSCO, GreenFILE-EBSCO, Web of Science and WHO-IRIS). Screening will be done independently by at least two reviewers, with disagreements resolve by consensus. We will extract data for the included studies using a standardised data extraction in an independent fashion, with disagreements resolved by consensus. We will conduct a narrative synthesis.

## Background

Globally, tobacco is the largest cause of preventable deaths, killing more than 8 million and many more suffer from other diseases conditions associated with it. India is the 2^nd^ largest producer and consumer of tobacco products and tobacco is consumed in myriad forms ranging from smoked products like cigarette and bidi, and plethora of smokeless products. However, unlike other countries, where cigarettes are predominant form of tobacco being consumed, bidi is the most commonly smoked product in India. Bidis are an indigenous smoking tobacco product, made by tobacco flakes rolled in tendu leaves and tied with a thread. They roughly contain 0.15–0.5-gram tobacco per stick which amounts to one-fifth to two-thirds of the amount of tobacco in cigarettes. However, the concentration of nicotine in bidis is higher than that in cigarettes and it delivers higher levels of tar compared to cigarettes its smoke leading to potentially greater health risk.

On analysing the constituents of bidi, the Ministry of Health and Family Welfare (MoHFW), Government of India (GoI) monograph concluded that the low porosity of the *bidi* leaf wrapper and low filtration of *bidi*s contribute to higher amounts of toxins released during *bidi* smoking^1^.

Evidence from epidemiological studies suggest that bidi smoking is associated with many health risks like severe respiratory impairment, increased risk of oral cancer and lung cancer, all-cause mortality, and adverse cardiorespiratory outcomes^2,3^. Apart from the adverse health outcomes among *bidi* smokers, bidi rollers are exposed to several occupational health hazards because of long working hours and unhealthy work environment^4^. Bidi manufacturing is an agro-forest based cottage industry and is generally rolled at home. These workers are exposed to nicotine, tar and other particles through skin leading to occupational health risks in not only themselves but also in their families and communities. Environmental effects of bidi manufacturing particularly in homes is also another cause of concern^5^. There is however no synthesises of the available evidence on the environmental risks and health hazards in bidi workers, their families, and communities to inform policy makers and other stakeholders. On request from WHO-India we sought to conduct a synthesis of evidence on this domain and develop a policy brief based on it.

## Methods

### Broad Approach

We will conduct a systematic scoping review of the available evidence on the topic of interest. The scoping approach is appropriate as the topic at hand is broad and there is no effectiveness or safety question at hand. Based on the scoping review, an evidence-informed policy brief will be created.

### Eligibility Criteria

We will include studies and reports, which meet the following criteria

- Population / problem – are on bidi workers (any role, function or type of enterprise), their families, communities in the working environment
- Interest area - are on any of the following:
  ∘ occupational health hazards of bidi workers
  ∘ health hazards in families and communities of bidi worker
  ∘ environmental risk of bidi manufacturing /rolling
- Setting – the study should be conducted in India
- Study design - presents any primary data (qualitative or quantitative) irrespective of study design or peer-review status

We will include only English language documents. We will not include animal or laboratory studies on effects of constitutes of bidi.

### Information sources

#### Electronic database search

The search strategy for PubMed has been developed and presented in **Appendix 1**. This will be adapted for other electronic databases.

We will search multiple indexed databases, from their date of inception

1. PubMed,
2. EMBASE+EMBASE Classic
3. CINAHL
4. Environment complete-EBSCO
5. GreenFILE – EBSCO
6. Web of Science
7. WHO-IRIS

All search strategies will be made available as appendix along with full report.

#### Other methods for searching

We will also use following additional modalities to identify potential studies from the grey literature

- Through expert contact and citation tracking (from amongst those identified to be included by other means).
- Hand searching websites of Ministry of Health and Ministry of labour of government of India, labour organisations, trade unions in India, and non-profit organisation working on tobacco control, or welfare of bidi workers in India. A full list of all websites searched will be presented with the full review.

### Screening process and Data Collection

Two reviewers will independently screen title and/or abstracts from electronic database search and, disagreement if any will be solved by consensus. Full text evaluation will be done by at least two reviewers. We will collect data as per a standardised data extraction form which will be piloted and iteratively developed.

### Synthesis of results

We will narratively synthesise the study without any additional quantitative analysis. Given the broad nature of question and substantial differences in context, narrative synthesis is the right approach for the process. Gaps in knowledge will be identified and presented. We will synthesise and report the occupational hazards, health risk and environmental risk of bidi workers, their families and communities into following sub-groups, if enough studies are available for each sub-type:

- Male and female bidi workers
- Child labourers in the bidi industry
- Pregnant and lactating bidi workers
- Full-time and part-time bidi workers
- Directly employed and contracted bidi workers
- Type of enterprise –
  ∘ Own Account Manufacturing Enterprises (OAME): A small manufacturing enterprise run in household of bidi workers, and that do not hire any workers on a regular basis (contracted out)
  ∘ Establishments: An enterprise which has at least one hired worker on a regular basis

## Supporting information

Appendix 1 : Search Strategy for Pubmed

## Data Availability

All data related to the protocol is contained in the manuscript or accompanying supplementary material

## Author contributions

Jyoti Tyagi: scoping to inform protocol; search strategy; design search strategy; first draft protocol

Deepti Beri: scoping to inform protocol, review and edit draft protocol with sufficient intellectual input

Soumyadeep Bhaumik: conceptualisation; methodology; first draft of protocol

Guarantors: all authors

