## Appendix 1 : Search Strategy for Pubmed for "Environmental risks and occupational health hazards of bidi workers and their communities in India: protocol for systematic scoping review"

| **No.** | **Search terms** |
| --- | --- |
| #1 | “bidi” OR bidis OR “beedi” OR beedis |
| #2 | India[Mesh] OR India* OR Andaman OR Nicobar OR Andhra OR Arunachal OR Assam OR Bihar OR Chandigarh OR Chhattisgarh OR “Dadra and Nagar Haveli” OR Daman OR Diu OR Delhi OR Goa OR Gujarat OR Haryana OR Himachal OR Jammu OR Kashmir OR Jharkhand OR Karnataka OR Kerala OR Lakshadweep OR “Madhya Pradesh” OR Maharashtra OR Manipur OR Meghalaya OR Mizoram OR Nagaland OR Orissa OR Odisha OR Pondicherry OR Punjab OR Rajasthan OR Sikkim OR “Tamil Nadu” OR Telangana OR Tripura OR “Uttar Pradesh” OR Uttarakhand OR “Bengal” |
| #3 | #1 AND #2 |
